# Jinhua Qinggan Granules for Nonhospitalized COVID-19 Patients: a Double-Blind, Placebo-Controlled, Randomized Controlled Trial

**DOI:** 10.1101/2022.05.16.22275074

**Authors:** Muhammad Raza Shah, Samreen Fatima, Sehrosh Naz Khan, Shafiullah, Gulshan Himani, Kelvin Wan, Timothy Lin, Johnson Y.N. Lau, Qingquan Liu, Dennis S.C. Lam

## Abstract

**Background:** Key findings from the World Health Organization Expert Meeting on Evaluation of Traditional Chinese Medicine in treating COVID-19 reported that TCMs are beneficial, particularly for mild-to-moderate cases. The efficacy of Jinhua Qinggan Granules (JHQG) in COVID-19 patients with mild symptoms has yet to be clearly defined.

**Methods:** We conducted a phase 2/3, double-blind, randomized, placebo-controlled trial to evaluate the efficacy and safety of treatment with JHQG in mild, nonhospitalized, laboratory-confirmed COVID-19 patients. Participants were randomly assigned to receive 5g/sacket of JHQG or placebo granules orally thrice daily for 10 days. The primary outcomes were the improvement in clinical symptoms and proportion tested negative on viral PCR after treatment. Secondary outcomes were the time to recovery from clinical symptoms and changes in white blood cells (WBC) and acute phase reactants (C-reactive protein (CRP) and ferritin) 10-15 days after treatment.

**Results:** A total of 300 patients were randomly assigned to receive JHQG (150 patients) and placebo (150 patients). Baseline characteristics were similar in the two groups. In the modified intention-to-treat analysis, JHQG showed greater clinical efficacy (82.67%) after 10 days of treatment compared with the placebo group (10.74%) (rate difference: 71.93%; 95% CI 64.09 - 79.76). The proportion of patients with a negative PCR after treatment were comparable (rate difference: -4.67%; 95% CI -15.76 - 6.42). While all changes in WBC, ferritin, and CRP levels showed a statistically significant decline in JHQG (P≤0.044) after treatment, but not the latter in placebo (P=0.077). The median time to recovery of COVID-19 related symptoms including cough, sputum, sore throat, dyspnea, headache, nasal obstruction, fatigue, and myalgia were shorter in the JHQG group compared to the placebo group (P<0.001 for all). 3 patients experienced mild to moderate adverse events during the treatment period in the JHQG group. Findings were similar between the modified intention-to-treat and the per-protocol analysis that included only patients who reported 100% adherence to the assigned regimen.

**Conclusions:** JHQG is a safe and effective TCM for the treatment of mild COVID-19 patients.

**Clinical Trial Registration:** The Trial was prospectively registered on www.clinicaltrials.gov with registration number: NCT04723524.

## 1 Introduction

The coronavirus disease 2019 (COVID-19) caused by the severe acute respiratory syndrome coronavirus 2 (SARS-CoV-2) came under the attention of the international medical community when China first notified the World Health Organization (WHO) of a pneumonia outbreak of then unknown etiology in December 2019 [1]. COVID-19 was subsequently declared by the WHO as a Public Health Emergency of International Concern and further as a pandemic as SARS-CoV-2 infections surged globally [2, 3]. With the advent of the highly contagious Omicron and potentially emerging novel variants, the COVID-19 pandemic remains a threat to public health worldwide [4].

As of 4 April 2022, COVID-19 has resulted in over 490 million cases and more than 6.1 million deaths globally [5]. Although various vaccines showed evidence of substantially reducing hospitalization and mortality, limited access and public hesitance to vaccination have hindered the attainment of herd immunity to halt the pandemic through vaccination [6, 7]. Specific populations of patients, in particular elderly patients and patients with chronic medical conditions (e.g., obesity, diabetes mellitus, malignancy, pulmonary diseases, cardiovascular diseases etc.) are at significantly heightened risk of progression to severe disease and mortality after SARS-CoV-2 infection [8, 9]. Therefore, a demand for anti-COVID-19 treatment options which can prevent the progression to severe disease and mortality exists. Such therapeutic agents should be readily available to be administered to patients with mild COVID-19 at disease onset to prevent subsequent progression.

Currently, oral antiviral agents available under emergency use authorization by the Food and Drug Administration for COVID-19 include molnupiravir and nirmatrelvir [10]. Recently, the WHO Expert Meeting on Evaluation of Traditional Chinese Medicine (TCM) in the Treatment of COVID-19 has concluded that traditional Chinese medicine is beneficial in mild-to-moderate COVID-19, as on add-on interventions to conventional treatment, TCM may shorten the time for viral clearance, resolution of clinical symptoms and length of hospital stay [11]. Various TCMs have been approved by China’s National Administration of TCM to manage COVID-19 [12]. In the *Diagnosis and Treatment Protocol for Novel Coronavirus Pneumonia (Trial Version 7)* released by China’s National Health Commission & National Administration of TCM, Jinhua Qinggan Granules (JHQG) was recommended as a treatment for fatigue and fever for COVID-19 patients during the medical observation period [13].

JHQG was previously proposed as a potential TCM for the treatment of influenza and has been shown to shorten the duration of fever and recovery time in influenza patients [14]. In recent studies of JHQG in COVID-19 patients, JHQG was reported to increase the viral clearance rate in COVID-19 patients as evidenced by an increased proportion of patients with negative nucleic acid tests after JHQG treatment [15]. It has further been studied in a randomized controlled trial (RCT) which demonstrated that COVID-19 patients treated with JHQG combined with Western medications (oseltamivir and arbidol) showed increased improvement in fever and fatigue compared with patients treated with the antivirals alone [16]. Nevertheless, the efficacy and safety of the independent use of JHQG in the treatment of COVID-19 patients for the prevention of disease progression remain to be elucidated. Therefore, this clinical trial was aimed to evaluate the efficacy and safety of JHQG granules in nonhospitalized Pakistani COVID-19 patients with mild disease.

## 2 Methods

This phase 2-3, double-blind, randomized, placebo-controlled clinical trial evaluated the efficacy and safety associated with the use of JHQG among nonhospitalized COVID-19 adult Pakistani patients with mild symptoms. This study was approved by the Institutional Ethics Committee of International Center for chemical and biological Sciences (ICCBS) and institutional review board of the Indus Hospital (Sector 39, Karachi, Sindh, Pakistan). The trial was prospectively registered on www.clinicaltrials.gov with registration number: NCT04723524. All study participants provided written informed consent before enrolling in the clinical trial.

### 2.1 Eligibility

Eligible patients to be enrolled in this study fulfilled all of the following inclusion criteria: (1) age range of 18-75 years; (2) Confirmed SARS-CoV-2 infection by real time reverse transcription polymerase chain reaction (RT-PCR); (3) Mild symptoms cases of grade 2 having any of COVID-19 related fever, sore throat, cough, headache, malaise, nausea, vomiting, diarrhea, muscle pain, or loss of taste and smell symptoms; and (4) capable of providing written informed consent. Patients were excluded if they had any of the following: (1) previous confirmed SARS-CoV-2 infection; (2) moderate or critical COVID-19 infection with (a) respiratory failure and requiring mechanical ventilation, (b) shock, or (c) other organ failure requiring intensive care unit (ICU) support; (3) severe primary health conditions associated with cardiovascular, cerebrovascular, pulmonary, hepatic, renal, endocrine and hematological diseases, hematopoietic system (above grade II of cardiac function; ALT & AST are 1.5 times higher than the normal value; Creatinine above the upper limit of normal value) and mental illness or serious diseases affecting their survival, such as cancer or AIDS; (4) administered other antiviral, antibiotics, cough relieving and antihistamine medications within 3 days prior to the visit (including β2 receptor agonists, anticholinergic agents, theophylline, glucocorticoids, cough expectorant and other TCM); (5) history of drug or food allergy; (6) pregnancy, lactating, or fertile women who were planning to conceive in 3 months; and (7) participated in another clinical study in the past 1 month.

### 2.2 Sample Size Calculation

The sample size was based on detecting a 20% difference in recovery from COVID-19 related symptoms. Defined by asymptomatic clinical state (40% with placebo vs 60% with TCM) at 90% power, 260 patients (130 per group) are required at p<0.05. A sample size of 300 patients was adopted to allow for approximately 15% loss to follow-up without compromising the statistical power.

### 2.3 Drug under Investigation

Jinhua Qinggan granules (JHQG; Chinese medicine Z20160001; Ju Xie Chang (Beijing) Pharmaceuticals Co., Ltd. (Jingjintang Kejiyuan Zhengzhong, Daxing, Beijing, China)) having Batch no. 20200601 containing 5g JHQG or placebo per sachets were used in this study. JHQG is synthesized from the two TCMs formulae Ma Xing Shi Gan Decoction and Yin Qiao San Decoction [17]. Packaging of the JHQG and placebo were the same and conformed to regulations of the Chinese Pharmacopoeia (Chinese Pharmacopoeia Commission, 2015) in terms of quality standards.

### 2.4 Patient Allocation and Assessments

Eligible patients were randomly assigned in a 1:1 ratio using a centralized, interactive-response technology system to receive either JHQG (5g/sachet) or matched placebo, administered thrice daily for 10 days. Patients and investigators in this trial were blinded to the treatment allocation until the completion of the study. The computer-generated random numbers were used to generate randomization sequence for allocation of participants to TCM or placebo. Data analysis was performed independently by professional statisticians to guarantee that all enrolled participants were evenly allocated to the JHQG or Placebo groups. The drug sequence was randomly assigned to the subjects according to the randomization list and each subject received the medication according to their randomization list. In-case of severe adverse events (SAEs) or other unwanted situations, urgent un-blinding permission was granted under principal investigator (PI). Eligible patients received JHQG or Placebo (5g/sachet) at an oral dose of 5 g (1 Sachet) three times a day after meal dissolved in boiled water for 10 days. The course of treatment was 10 days, and the visit at 10-15^th^ day of treatment was set as follow-up. Patients were assessed by a seven-category ordinal scale (**Table S1;** supplementary File) and clinical signs and symptoms on day 1 and 10-15^th^ day. All patients were reviewed on daily basis by investigators via telephone calls for medication consumption, health status and patient diary record through day 10. The detailed assessment schedule is outlined in **Table S1**.

### 2.5 Study Endpoints

Eligible patients were assessed using a seven-category ordinal scale and clinical signs and symptoms on 1^st^ and 10-15^th^ day of the trail period were recorded (**Table S2;** supplementary File). All effectiveness and safety inspection items were done once before the trial and once at follow-up i.e., 10 -15^th^ day. Demographics, vital signs, clinical symptoms, medication status, adverse events were recorded to evaluate the participants’ degree of COVID-19 related symptom improvement (efficacy) according to the schedule given in **Table S1** (supplementary File). Routine laboratory blood and urinary tests, electrocardiogram (ECG), serum electrolytes, liver function, renal function, creatine phosphokinase, SARS-CoV-2 RT-PCR, C-reactive protein (CRP), ferritin, and chest X-ray examination were performed at 1^st^ and 10-15^th^ day of the trial to detect abnormal changes and assess the safety of the drug.

#### 2.5.1 Efficacy Endpoints

The primary efficacy endpoints of the study were (i) the improvement in clinical signs and symptoms based on the seven-category ordinal scale (**Table S1**) obtained on day 0 and 10-15 days after treatment initiation; and (ii) the proportion of patients who tested negative to nasopharyngeal SARS-CoV-2 test on RT-PCR 10 to 15 days after treatment. Secondary efficacy endpoints were based on the documentation of body temperature; change in white blood cells (WBC), CRP, and ferritin levels; change in radiographic (chest X-ray) findings; time to recovery of individual symptom; and quality of life assessment (given in supplimentary file) at day 0 and 10-15 days after treatment initiation. The primary symptoms for improvement assessment were cough and fever while sputum, sore throat, dyspnea, headache, nasal obstruction, myalgia, runny nose, chest pain, and diarrhea were secondary symptoms. The recovery time from individual COVID-19 related symptoms i.e. cough, sputum, sore throat, dyspnea, headache, nasal obstruction, myalgia, runny nose, chest pain, and diarrhea was analyzed using Kaplan-Meier to calculate the median time followed by Log-rank to check the differences between the JHQG and placebo groups. Clinical efficacy of JHQG was judged based on the effectiveness of the drug in terms of before and after treatment scores of main symptoms and secondary symptoms as Remarkable effective (Cough and fever are remarkably improved; Effective (Cough and fever are improved) and Ineffective (No improvement of cough). The curative effect of JHQG post treatment was analyzed through curative index analysis as Clinically cured (symptom grade decreased ≥90%), Remarkable effective (symptom grade decreased ≥70% and <90%), Effective (symptom grade decreases ≥30% and <70%), Ineffective (symptom grade decreased ≥0% and <30%) and Worsened (symptom grade increased <0%). The radiologist who analysed and graded te chest X-ray of the patients were kept blinded.

#### 2.5.2 Safety

Safety endpoints included various clinical investigation results obtained from patients during the study period: blood pressure, heart rate, and respiration rate), ECG, chest X-ray at screening, and follow-up visits. Routine blood tests, urinalysis, serum electrolytes, liver function tests (ALT, AST, TBIL, AKP, γ-GT), and renal function tests (BUN, Cr) of patients in both groups at day 0 and 10-15^th^ day after treatment were performed. Medication compliance and adverse events were assessed for all patients on follow-up (10-15^th^ day). The rate of disease aggravation after 10 days of treatment was also evaluated. All patients actively recorded any adverse events (AEs) potentially related to treatment with details including their occurrence, remission, and severity in patient diaries which were transcribed into detailed case report forms for investigators’ review. The AEs were classified into Mild AEs (asymptomatic or mild symptoms with no intervention indicated), moderate AEs (minimal, local or noninvasive intervention indicated) or severe AEs (disabling; hospitalization or prolongation of hospitalization indicated; medically significant but not immediately life-threatening).

### 2.6 Statistical Analysis

The full analysis set (FAS) included subjects as close as possible to intention-to-treat principle and was used to analyze primary and secondary endpoints. The last observation carried forward method (LOCF) was used to estimate the missing values of the primary endpoints. The Per Protocol Set (PPS) consisted of all the subjects that complied with the study protocol, drug compliance (80 - 120%), and had complete records required in the case report form (CRF). PPS was used for primary and secondary efficacy endpoints analysis. Statistical analysis was performed using SPSS (version 23.0; IBM, NY, USA). Descriptive statistics were reported as proportions, mean ± standard deviation (SD), and median (interquartile range [IQR]) where appropriate. The comparison of continuous variables was made using t-test/rank-sum test and chi-square test or Fisher’s probability exact test was used for the comparison of discrete and categorical variables. All statistical inferences used two-sided tests, with a statistically significant test level of 0.05. Efficacy analysis was applied to both FAS and PPS while safety analysis was applied to Safety Set (SS) consisting of all randomized subjects that have used the test drug at least once and have at least one safety assessment record.

## 3 Results

### 3.1 Patient sample and characteristics

A total of 402 RT-PCR confirmed COVID-19 patients were identified from September 22, 2020 to August 23, 2021 at Indus Hospital (Plot C-76, Korangi Crossing, Karachi, Sindh) Pakistan. Eligible patients (n=300) were recruited and randomly allocated in 1:1 ratio into the JHQG (n= 150) and placebo (n=150, Control) groups as shown in **Figure 1**. The enrolled patients were from different ethnicities including 174 Urdu (58%), 12 Pashtuns (4.0%), 6 Baluchis (2.0%), 6 Gilgiti (2.0%), 56 Punjabi (18.66%), 15 Sindhi (5.0%), and 31 others (10.33%). Two patients were eliminated from the study due to concomitant medication use and 42 patients withdrew consent after randomization. Thus, a total of 256 patients completed the study and were included in final analysis (PPS analysis). Both groups had no statistical differences (P>0.05) in terms of demographic characteristics, concomitant medications, and past medical history (**Table 1**).

**Table 1.**
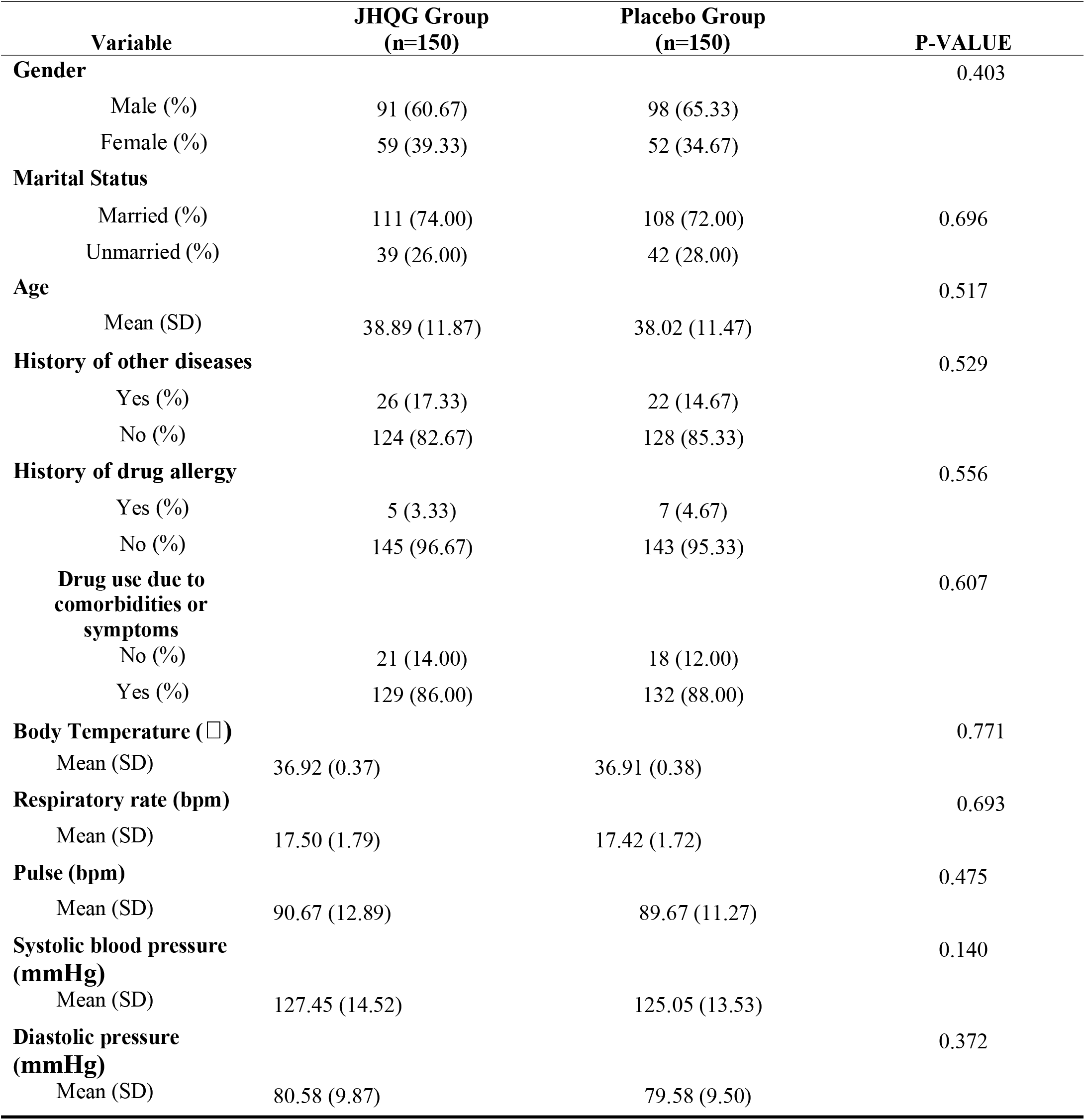
Demographics and Baseline Characteristics of the subjects in the trial.

**Figure 1.**
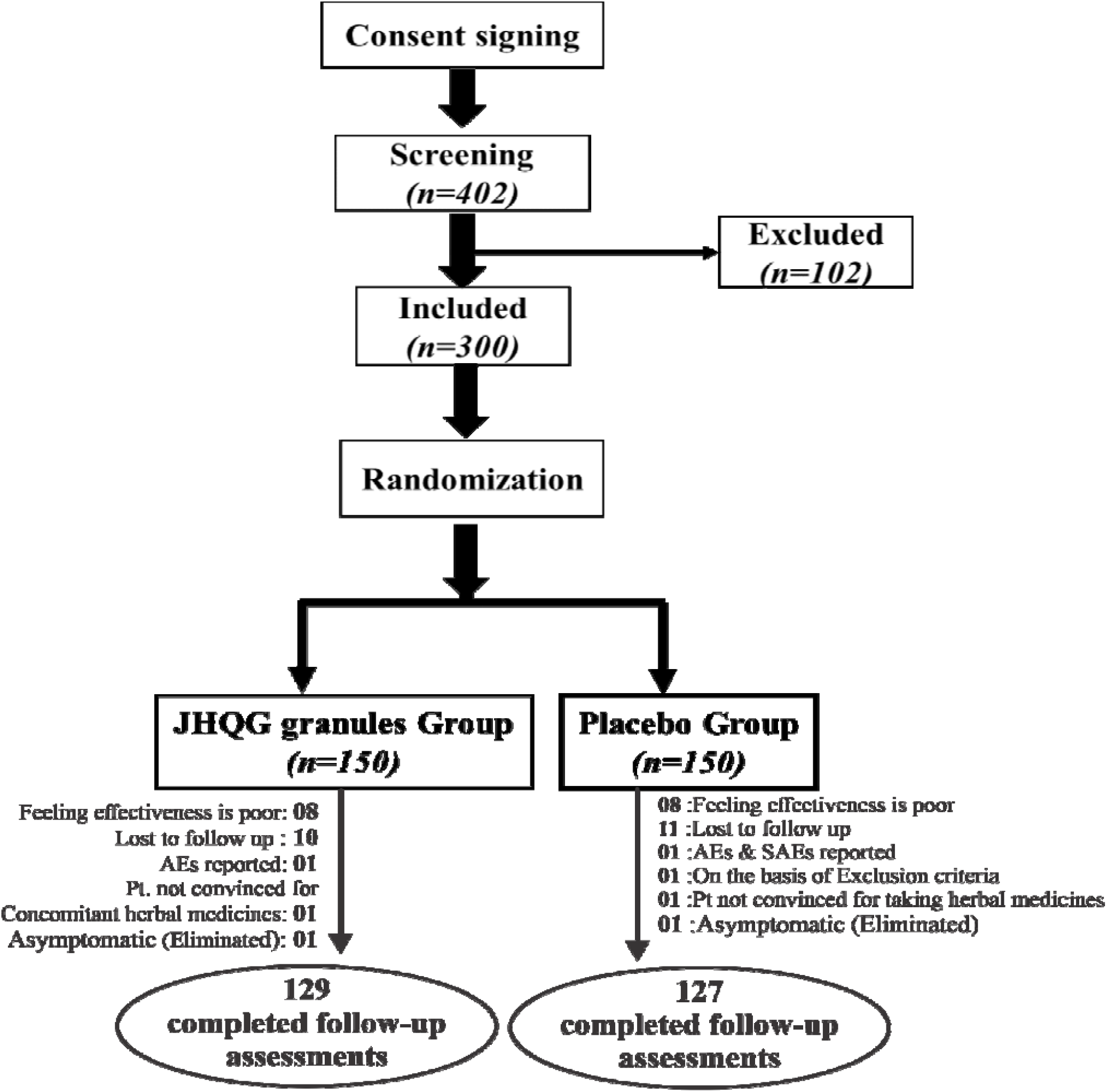
Flowchart of screening, randomization, and treatment of subjects.

### 3.2 Primary Efficacy End Points

Based on FAS and PPS, each subject’s score of main symptoms (MS) and secondary symptoms (SS) as well as total scores (TS) were described and compared at baseline and at follow-up for JHQG and placebo group. The inter-group difference had no statistical significance (P>0.05) at baseline (**Table 2**). Clinical efficacy after treatment was analyzed for each group. FAS showed that the clinical efficacy was 82.67% and 10.74% for the JHQG and placebo groups, respectively, and the rate difference was 71.93% (95% CI 64.09 - 79.76), suggesting that JHQG was superior to placebo (**Table 3**). The PPS analysis showed that the clinical efficacy was 95.35% and 12.60% for the JHQG and placebo groups, respectively, and the rate difference was 82.75 (95% CI 75.93 -89.57) (**Table 3**). Both the FAS and PPS analyses for clinical efficacy comparison between the JHQG and placebo groups showed statistically significant differences (P<0.001 for both) indicating that JHQG was superior to placebo. In FAS, the post-treatment SARS-CoV-2 negative test rate was 38.00% and 42.67% in the JHQG and placebo groups respectively, with a rate difference of -4.67 (95% CI -15.76 - 6.42). In PPS, the post-treatment SARS-CoV-2 negative test rate was 44.19% and 44.61% in the JHQG and placebo groups, respectively, with a rate difference of -5.42 (95% CI -17.63 to 6.79), and both analyses failed to reach statistically significant outcomes (p>0.05, **Table 4**).

**Table 2.** Symptom Scores(FAS)and (PPS) of the patients.

| **Symptom Score ( PPS )** |  |  |  |
| --- | --- | --- | --- |
| **Variable** | **JHQG Group**<br>**(n=150)** | **Placebo Group**<br>**(n=150)** | **P-VALUE** |
| **MS Score** |  |  | 0.782 |
| Mean (SD) | 2.35 (0.83) | 2.32 (0.84) |  |
| Min, Max | 0.00, 6.00 | 0.00, 6.00 |  |
| **SS Score** |  |  | 0.848 |
| Mean (SD) | 8.49 (4.47) | 8.59 (3.94) |  |
| Min, Max | 0.00, 22.00 | 0.00, 18.00 |  |
| **TS Score** |  |  | 0.896 |
| Mean (SD) | 10.84 (4.65) | 10.91 (4.19) |  |
| Min, Max | 0.00, 24.00 | 0.00, 22.00 |  |
| **Symptom Score ( PPS )** |  |  |  |
| **Variable** | **JHQG Group**<br>**(n=129)** | **Placebo Group**<br>**(n=127)** | **P-VALUE** |
| **MS Score** |  |  | 0.579 |
| Mean (SD) | 2.31 (0.73) | 2.36 (0.77) |  |
| Min, Max | 2.00, 4.00 | 2.00, 4.00 |  |
| **SS Score** |  |  | 0.750 |
| Mean (SD) | 8.45 (4.34) | 8.61 (3.88) |  |
| Min, Max | 0.00, 22.00 | 0.00, 18.00 |  |
| **TS Score** |  |  | 0.685 |
| Mean (SD) | 10.76 (4.43) | 10.98 (4.09) |  |
| Min, Max | 2.00, 24.00 | 2.00, 20.00 |  |

MS= Main symptoms, SS= Secondary symptoms, TS= Total symptoms

**Table 3.**
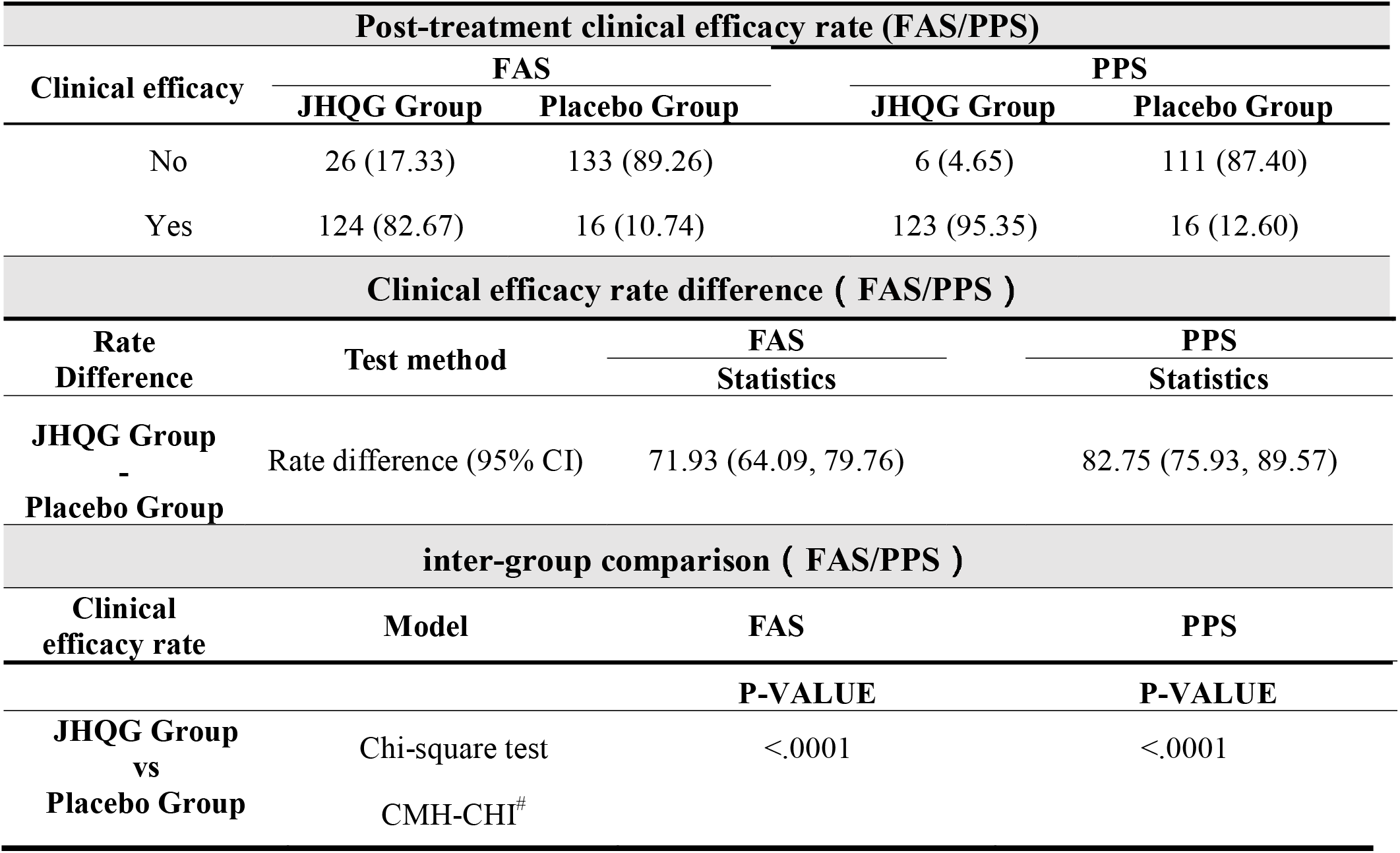
Post-treatment clinical efficacy rate (FAS/PPS), rate difference(FAS/PPS)and inter-group comparison.

| Post-treatment clinical efficacy rate (FAS/PPS) |  |  |  |  |
| --- | --- | --- | --- | --- |
| Clinical efficacy | FAS |  | PPS |  |
|  | JHQG Group | Placebo Group | JHQG Group | Placebo Group |
| No | 26 (17.33) | 133 (89.26) | 6 (4.65) | 111 (87.40) |
| Yes | 124 (82.67) | 16 (10.74) | 123 (95.35) | 16 (12.60) |
| Clinical efficacy rate difference ( FAS/PPS ) |  |  |  |  |
| Rate Difference | Test method | FAS Statistics | PPS Statistics |  |
| JHQG Group - Placebo Group | Rate difference (95% CI) | 71.93 (64.09, 79.76) | 82.75 (75.93, 89.57) |  |
| inter-group comparison ( FAS/PPS ) |  |  |  |  |
| Clinical efficacy rate | Model | FAS | PPS |  |
|  |  | P-VALUE | P-VALUE |  |
| JHQG Group vs Placebo Group | Chi-square test | <.0001 | <.0001 |  |
|  | CMH-CHI<sup>#</sup> |  |  |  |

**Table 4.**
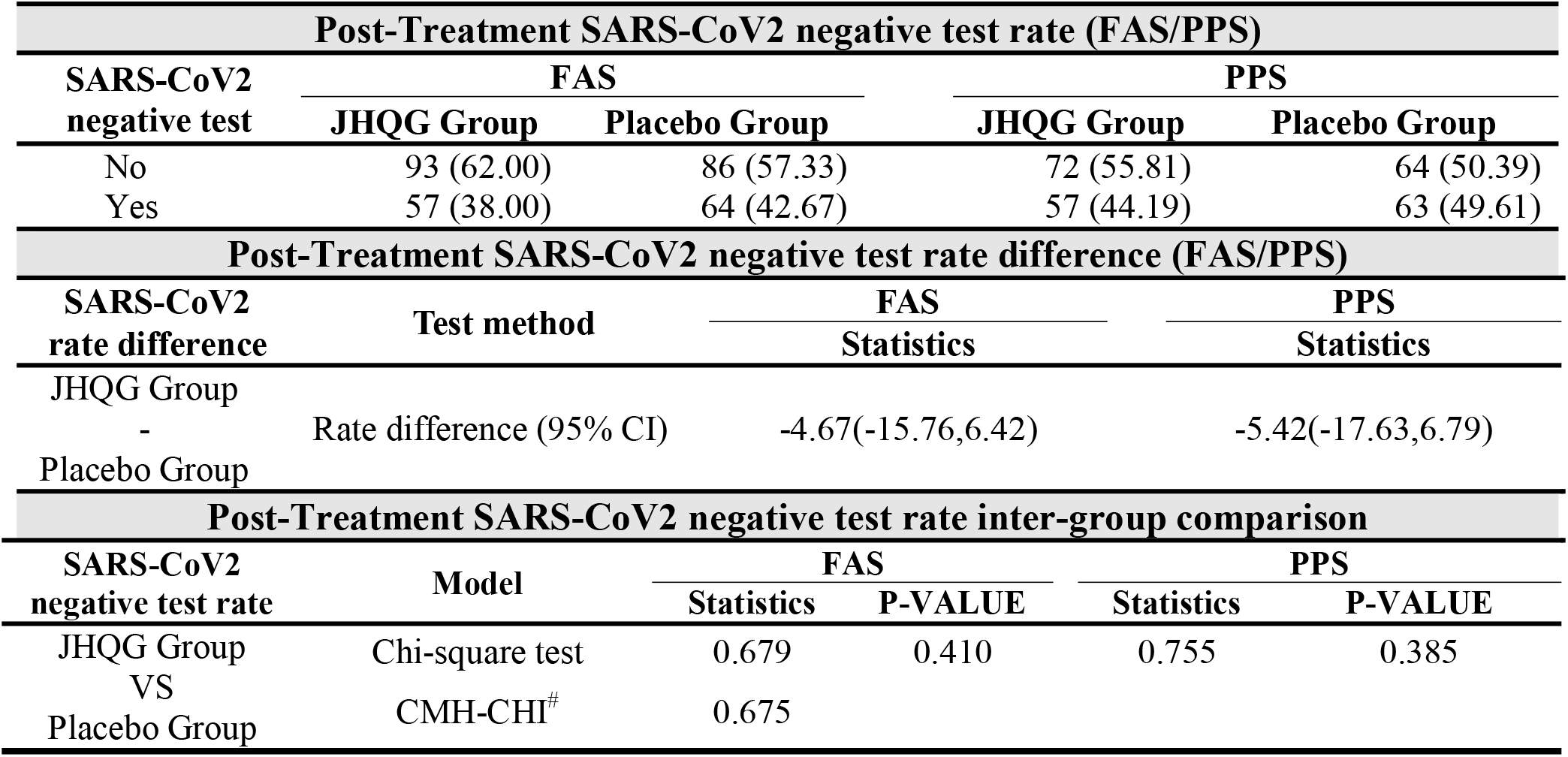
Post-Treatment SARS-CoV2 negative test rate (FAS/PPS), Test rate difference and Inter group comparison (FAS/PPS).

### 3.3 Secondary Efficacy End Points

The time of defervescence for JHQG was 2.00 days while it was 2.5 days for placebo (P>0.05). Inter-group comparison of the rate of clinically cured patients, remarkable effective rate, effective rate, ineffective rate, and worsened rate showed statistically significant differences in both FAS and PPS analysis of Curative index (P<0.001, **Table 5**). In the FAS analysis, the rate of clinically cured patients, remarkable effective rate, effective rate, ineffective rate, and worsened rate were 36.66%, 22.00%, 24.00%, 16.67% and 0.67% respectively in the JHQG arm, and 1.34%, 0.00%, 9.39%, 82.55%, and 6.71% respectively in the placebo arm. Inter-group comparisons showed statistically significant results (P<0.001). Similarly, in the PPS analysis, the rate of clinically cured patients, remarkable effective rate, effective rate, ineffective rate and worsened rate were 42.63, 25.58, 27.13, 3.87, and 0.77%, respectively in the JHQG arm, and 1.57, 0.00, 11.02, 79.52, and 7.87%, respectively in the placebo arm (P<0.001).

**Table 5.** Curative index analysis(FAS/PPS)

| Variable | JHQG Group | Placebo Group | P-VALUE |
| --- | --- | --- | --- |
| **FAS** |  |  | <0.001 |
| Worsened (%) | 1 (0.67) | 10 (6.66) |  |
| Ineffective (%) | 25 (16.67) | 123 (82.55) |  |
| Effective (%) | 36 (24.00) | 14 (9.39) |  |
| Remarkable E. (%) | 33 (22.00) | 0 (0.00) |  |
| Clinically Cured (%) | 55 (36.66) | 2 (1.34) |  |
| **PPS** |  |  | <0.001 |
| Worsened (%) | 1 (0.77) | 10 (7.87) |  |
| Ineffective (%) | 5 (3.87) | 101 (79.52) |  |
| Effective (%) | 35 (27.13) | 14 (11.02) |  |
| Remarkable E. (%) | 33 (25.58) | 0 (0.00) |  |
| Clinically Cured (%) | 55 (42.63) | 2 (1.57) |  |

Results of the QoL questionnaire showed that the QoL of patients in the JHQG group improved (1.96 ± 0.75) in comparison with the placebo group (0.44 ± 0.65) after treatment (FAS analysis, P<0.001). Similarly, in PPS analysis, results of the QoL questionnaire showed that the QoL of patients in the JHQG group improved (1.97 ± 0.75) in comparison with the placebo group (0.44 ± 0.65) (P<0.001) (**Table 6**).

**Table 6.** Change in Quality of Life (QoL) Questionnaire (FAS/PPS).

| Variable | JHQG Group | Placebo Group | P-VALUE |
| --- | --- | --- | --- |
| **FAS** |  |  |  |
| Before Treatment |  |  | <0.0001 |
| N (N miss) | 150 (0) | 150 (0) |  |
| Mean (SD) | 8.9277 (0.9695) | 9.8140 (0.8287) |  |
| After-Treatment |  |  | <0.0001 |
| N (N miss) | 130 (20) | 128 (22) |  |
| Mean (SD) | 10.7215 (0.6763) | 10.2743 (0.8489) |  |
| **PPS** |  |  |  |
| Before treatment |  |  | <0.0001 |
| N (N miss) | 129 (0) | 127 (0) |  |
| Mean (SD) | 8.7574 (0.8556) | 9.8334 (0.8568) |  |
| Post-Treatment |  |  | <0.0001 |
| N (N miss) | 129 (0) | 127 (0) |  |
| Mean (SD) | 10.7230 (0.6788) | 10.2687 (0.8499) |  |
| Before-After Treatment |  |  | <0.0001 |
| N (N miss) | 129 (0) | 127 (0) |  |
| Mean (SD) | 1.97 (0.75) | 0.44 (0.65) |  |

The change in radiographic findings in chest X-rays after treatment was also compared between the JHQG and placebo groups. In the FAS analysis, 12.5% of chest X-rays of patients in the JHQG group showed improvement 8.59% showed evidence of worsening after treatment, while 11.90% and 7.14% showed improvement and evidence of worsening after treatment, respectively, in the placebo group, with no statistically significant differences (P>0.05) between the 2 groups. Similarly, in the PPS analysis, 12.6% and 8.66% of chest X-rays of patients in the JHQG group showed improvement and evidence of worsening after treatment, respectively, while 11.90% and 7.14% showed improvement and evidence of worsening after treatment, respectively, in the placebo group, with no statistically significant differences (P>0.05) between the 2 groups (**Table 7**).

**Table 7.** Change in radiographic findings of the lungs (FAS/PPS).

| Variable | JHQG Group<br>n(%) | Placebo Group<br>n(%) | P-VALUE |
| --- | --- | --- | --- |
| **FAS** |  |  | 0.887 |
| Improvement | 16 (12.50) | 15 (11.90) |  |
| No change | 101 (78.91) | 102 (80.95) |  |
| Worsened | 11 (8.59) | 9 (7.14) |  |
| **PPS** |  |  | 0.887 |
| Improvement | 16 (12.60) | 15 (12.00) |  |
| No change | 100 (78.74) | 101 (80.80) |  |
| Worsened | 11 (8.66) | 9 (7.20) |  |

The incidence of adverse events in the JHQG and placebo groups showed no statistically significant differences (P>0.05, **Table 8**). In the JHQG group, 3 patients (2.0%) experienced adverse events. In the placebo group, 4 patients (2.67%) experienced adverse events. 2 patients and 4 incidences in the placebo group that had adverse events leading to withdrawal from study with 1.33% rate of incidence; while JHQG group had 1 patient and 2 incidences of adverse events that lead to withdrawal of consent with a 0.67% rate of incidence. In addition, 2 subjects (1.33%) withdrew consent due to adverse reactions in the placebo group, while no patient withdrew consent due to adverse reactions in JHQG group.

**Table 8.** Adverse events / reactions and their severity levels observed during the trial (SS).

|  | JHQG Group |  |  | Placebo Group |  |  | P-VALUE |
| --- | --- | --- | --- | --- | --- | --- | --- |
|  | Inc. | Sub. | Rate (%) | Inc. | Sub. | Rate (%) |  |
| **Adverse events** | **6** | **3** | **2.00** | **8** | **4** | **2.67** | **1.0000** |
| Mild | 4 | 2 | 1.33 | 2 | 1 | 0.67 | - |
| Moderate | 2 | 1 | 0.67 | 6 | 3 | 2.00 | - |
| Severe | 0 | 0 | 0.00 | 0 | 0 | 0.00 | - |
| **Adverse reactions** | **0** | **0** | **0.00** | **5** | **3** | **2.00** | **0.2475** |
| Mild | 0 | 0 | 0.00 | 2 | 1 | 0.67 | - |
| Moderate | 0 | 0 | 0.00 | 3 | 2 | 1.33 | - |
| Severe | 0 | 0 | 0.00 | 0 | 0 | 0.00 | - |
| *Adverse events that lead to withdrawal* | 2 | 1 | 0.67 | 4 | 2 | 1.33 | 1.0000 |
| *Adverse reactions that lead to withdrawal* | 0 | 0 | 0.00 | 4 | 2 | 1.33 | 0.4983 |

Inc.= incidences, Sub. = subjects.

The recovery time for the JHQG group from cough, sputum, sore throat, dyspnea, headache, nasal obstruction, fatigue, and myalgia symptoms was shorter than the placebo group (P<0.001). The median time (days) for recovery from cough, sputum, and sore throat symptoms JHQG was 6 days (for all) in the JHQG group and more than 11 days in the placebo group. The median time for recovery from fatigue was 7 days in the JHQG group and more than 11 days in the placebo group (P<0.001). There was no statistically significant differences between the two groups in the recovery time from runny nose, chest pain and diarrhea (P>0.05) (**Table 9**).

**Table 9.**
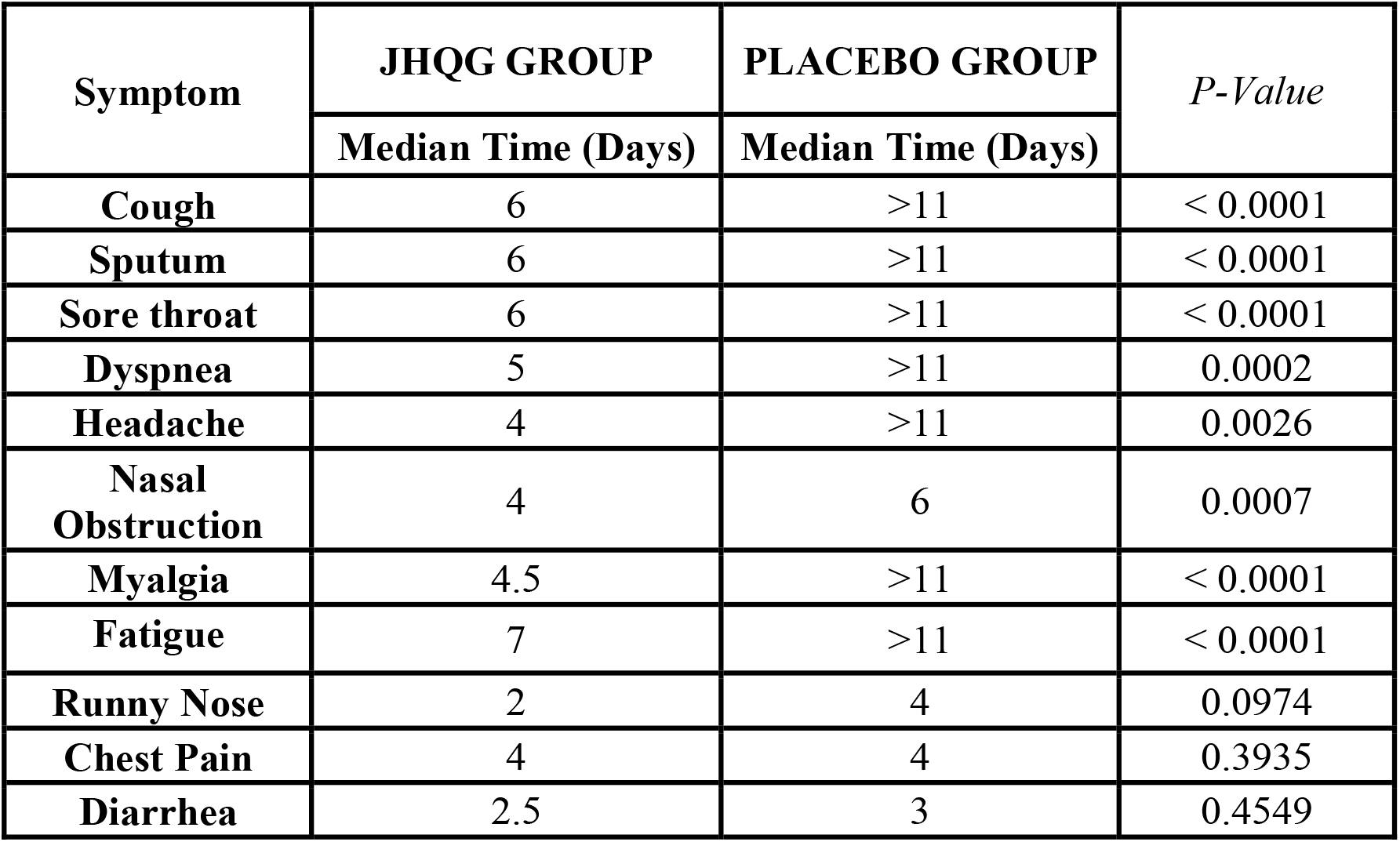
Comparison of the Recovery time for individual symptom in JHQG and Placebo Group.

| Symptom | JHQG GROUP | PLACEBO GROUP | *P-Value* |
| --- | --- | --- | --- |
|  | Median Time (Days) | Median Time (Days) |  |
| Cough | 6 | >11 | < 0.0001 |
| Sputum | 6 | >11 | < 0.0001 |
| Sore throat | 6 | >11 | < 0.0001 |
| Dyspnea | 5 | >11 | 0.0002 |
| Headache | 4 | >11 | 0.0026 |
| Nasal Obstruction | 4 | 6 | 0.0007 |
| Myalgia | 4.5 | >11 | < 0.0001 |
| Fatigue | 7 | >11 | < 0.0001 |
| Runny Nose | 2 | 4 | 0.0974 |
| Chest Pain | 4 | 4 | 0.3935 |
| Diarrhea | 2.5 | 3 | 0.4549 |

The changes in WBC, CRP, and ferritin levels were analyzed before and after treatment. In the FAS analysis, there were statistically significant intra-group differences for WBC levels in both the JHQG and placebo groups before and after treatment (P<0.001). The intra-group difference in CRP levels the JHQG group showed statistical significance (P=0.044) while there was no statistically significant difference in the placebo group (P=0.077). The intra-group difference in ferritin levels in both the JHQG and placebo groups showed statistically significant differences (P <0.001). Similar results were observed in the PPS analysis (**Table 10**). JHQG showed no statistically significant effects on routine blood tests, urinalysis, serum electrolytes, liver function tests (ALT, AST, TBIL, AKP, γ-GT), renal function tests (BUN, Cr) and ECGs of patients (Supplementary **Table S3 and S4**).

**Table 10.**
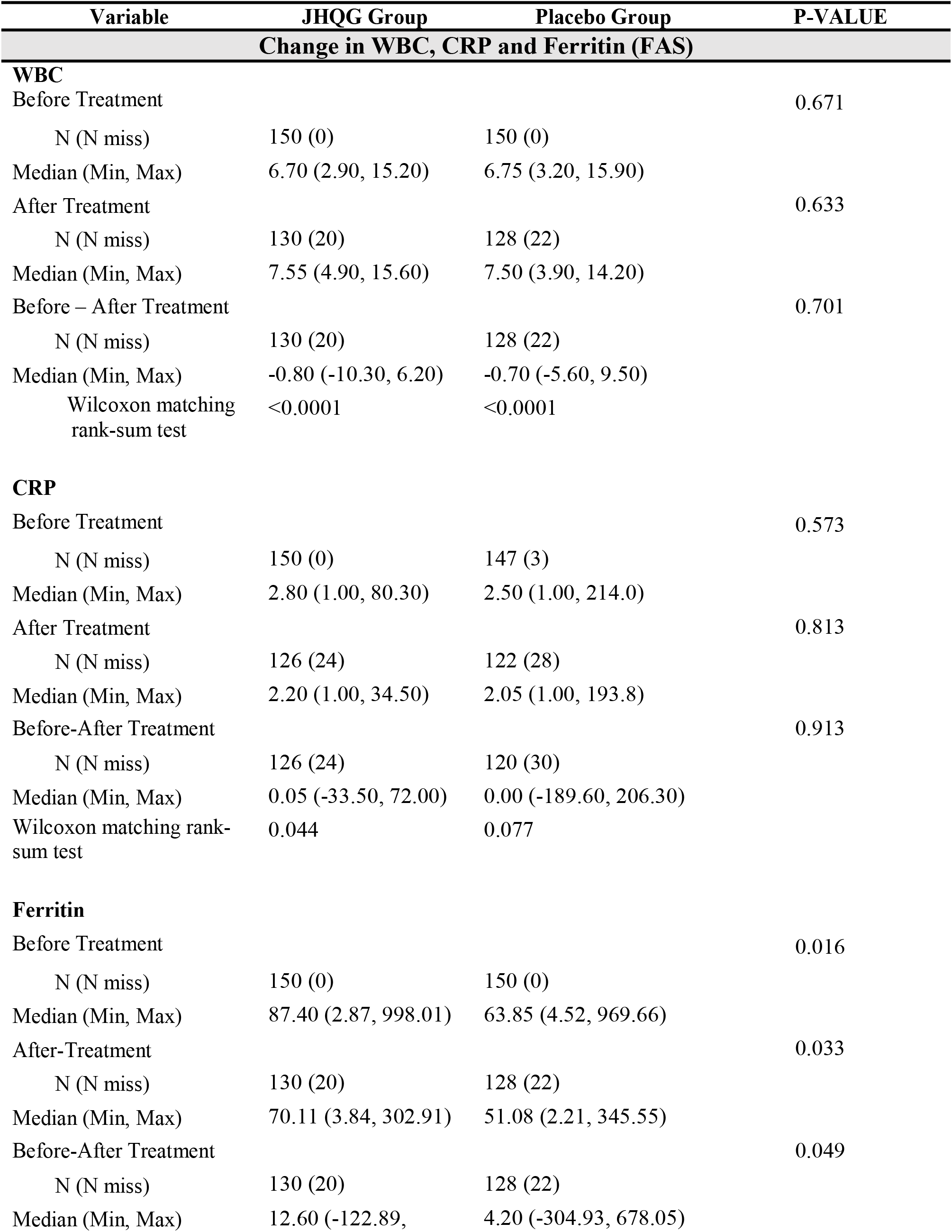
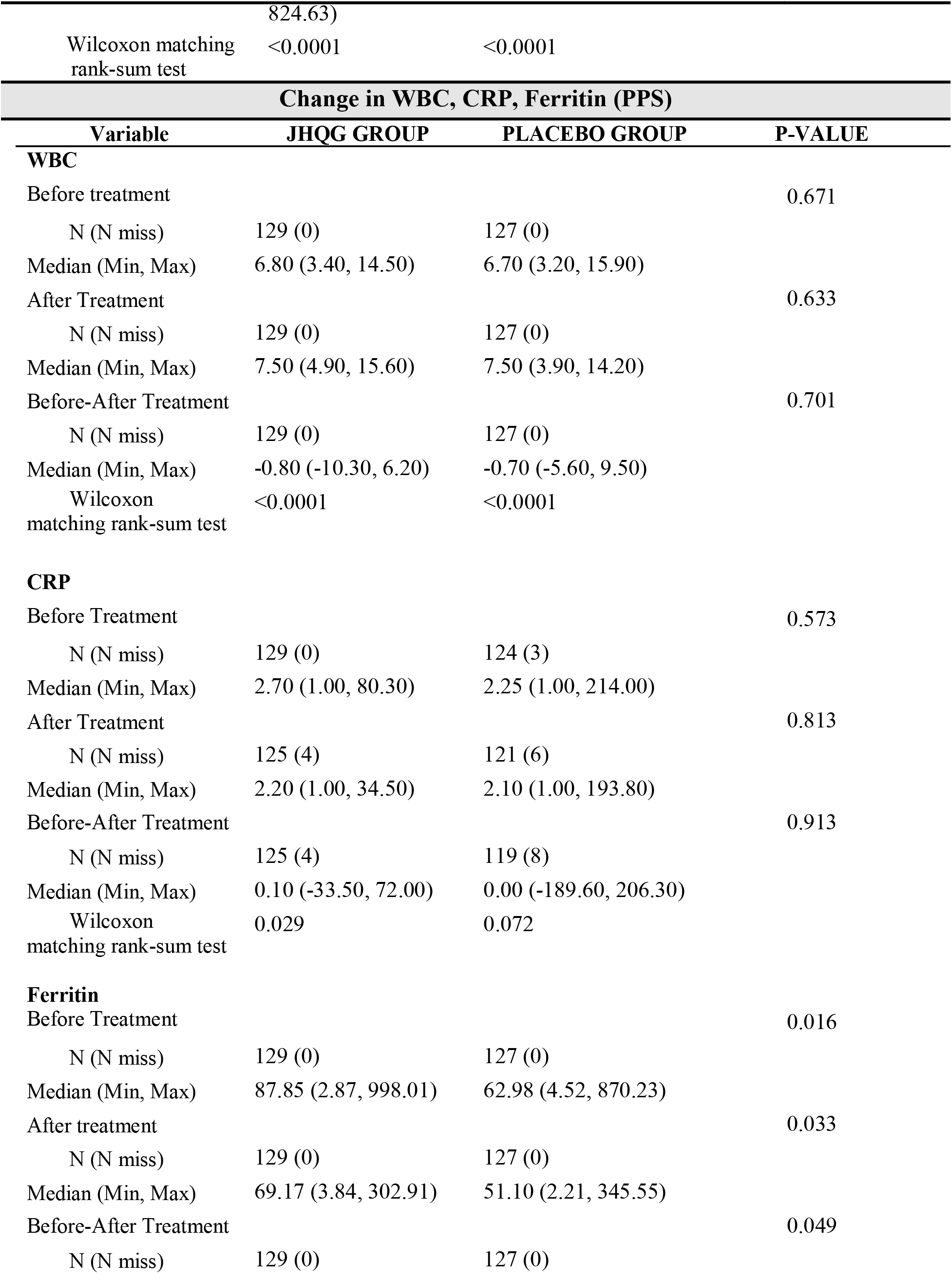
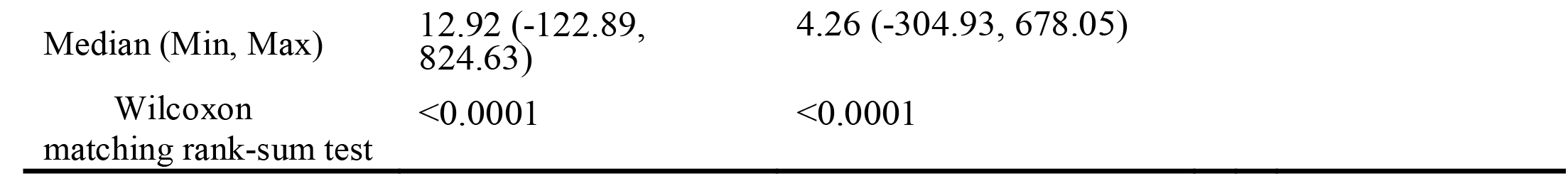
Change in WBC, CRP and Ferritin (FAS/PPS)

## 4 Discussion

In this study, we demonstrated that JHQG was a safe and effective TCM for the treatment of COVID-19 patients with mild symptoms. The data showed that JHQG (5g/sachet) administered orally to patients three times a day for 10 days achieved significant clinical efficacy in the treatment of COVID-19, reduced post-treatment WBC and acute phase reactant levels, shortened time of recovery for COVID-19 related symptoms, and improved QoL.

The Chinese National Health Commission & State Administration of Traditional Chinese Medicine has recommended JHQG to treat fatigue and fever in COVID-19 patients during the medical observation period [18]. In the 2009 H1N1 flu, studies have shown that the combined treatment of JHQG and oseltamivir can shorten the duration of fever [19, 20]. JHQG has shown a significant effect on treating mild COVID-19 related symptoms by shortening the period of fever, reducing inflammation, and improving symptoms [21-23]. JHQG is synthesized from Maxingshigan Decoction and Yinqiao Powder, and have the effects of “soothing wind,” ventilating the lungs, and clearing away heat and toxic materials [23].

A total of 256 patients, including 129 patients in the JHQG group and 127 patients in the placebo group completed the study. A total of 44 subjects dropped out (21 in the JHQG group and 23 in the Placebo group), causing 14.66% dropout rate. The basic reason for dropout was that the subjects were unable or unwilling to continue the clinical trial and voluntarily requested to withdraw. The JHQG group showed greater clinical efficacy (82.67 %) after 10 days of treatment compared with placebo (10.74%). The recovery time for JHQG group from cough, sputum, sore throat, dyspnea, headache, nasal obstruction, fatigue and myalgia symptoms was shorter as compared to placebo group (6 days vs >11 days; P<0.05). A previous study reported that JHQG could shorten the duration of fever, reduce the use of antibiotics and alleviate respiratory symptoms in patients with influenza A H1N1 [24]. In a recent RCT, JHQG granules combined with western medicine relieved the clinical symptoms of fever and poor appetite in COVID-19 patients and reduced the use of antibiotics to a certain extent [23]. It was suggested that certain compounds in JHQG could bind to specific target proteins and inhibit the activity of SARS-CoV-2 as revealed by high-throughput molecular docking and network pharmacology studies [25]. Various active ingredients in JHQG, including Quercetin and kaempferol, are hypothesized to target AEC2 and 3CL protein, inhibiting inflammatory mediators, eliminating free radicals, and regulating immunity [26]. These proposed mechanisms could explain the shortened recovery time of COVID-19 symptoms secondary to the host inflammatory response after infection.

The post-treatment SARS-CoV2 negative test rate for JHQG and placebo groups was 38.00% and 42.67% respectively. Though there was no statistically significant difference in SARS-CoV2 negative test rate in both groups (P>0.05), it was higher (38%) than a previous study (8.3%) [23]. Changes in WBC and CRP levels showed intra-group statistically significant differences for JHQG group. The ferritin intra-group differences after treatment for JHQG and Placebo groups showed statistically significant differences. The main chemical constituents of JHQG granules explored via modern pharmacological approach include stigmasterol, kaempferol, and quercetin, which possess anti-inflammatory, immunomodulatory, and antiviral effects [27]. In total, 10 adverse events/reactions were observed during the trial. Overall, JHQG was well tolerated and only 3 patients in the treatment group experienced mild to moderate adverse events. JHQG also showed no clinically significant effects on routine blood tests, urinalysis, serum electrolytes, liver function tests, renal function tests (BUN, Cr), and ECGs. These findings provide evidence to support that JHQG is a safe TCM among mild Covid-19 infection cases.

To our knowledge, this is the first RCT to evaluate the clinical efficacy and safety of JHQG in the treatment of laboratory-confirmed nonhospitalized COVID-19 patients. Various limitations of this trial should be noted. The basic reason for dropout was that the subjects were unable or unwilling to continue the clinical trial and voluntarily requested to withdraw The study included only COVID-19 patients of Pakistani race, and may limit the geographic generalizability of the findings. This study also excluded patients with severe underlying medical conditions, who are at particularly heightened risk of COVID-19 disease progression. Future studies of JHQG in COVID-19 shall focus on evaluating the clinical efficacy and safety of this TCM in such group of patients. In conclusion, our data show that JHQG is a safe and effective treatment for COVID-19 patients with mild symptoms.

## Data Availability

All data produced in the present study are available upon reasonable request to the authors.

## Conflict of interests

The authors declare that they have no conflict of interests.

## Funding

This Clinical Trial was funded by Ju Xie Chang (Beijing) Pharmaceuticals Co., Ltd. Jingjintang Kejiyuan Zhengzhong Street No.# 01, Daxing district, Beijing 102606, China.

## Ethical Statement

This study was conducted according to the ethical principles that have their origin in Declaration of Helsinki. The study protocol was reviewed and approved by the Institutional ethics committee of the International Center for Chemical and Biological Sciences (ICCBS), University of Karachi, Institutional Review Board of the Indus hospital (Sector 39, Karachi, Sindh, Pakistan) and National Bioethics committee (NBC) Pakistan. All study participants provided written informed consent before enrolling them in the clinical trial.

## Authors Contribution

RS: Principal investigator of the trial, Trail management; SF: Clinical Research Coordination; SK: Clinical Research Coordination, SU: Management of Trial and Regulatory affiars, GH: Clinical investigator; KW: Initial Draft preparation, TL: Data curation; JL: reviewing the Data; QL: Reviewing the manusctipt, DL: Finalized the data and manuscript.

## Notes

### Competing Interest Statement

The authors have declared no competing interest.

### Clinical Trial

NCT04723524

### Author Declarations

This study was approved by the Institutional Ethics Committee of International Center for chemical and biological Sciences (ICCBS) and institutional review board of the Indus Hospital (Sector 39, Karachi, Sindh, Pakistan).

