## Supplementary File for "Jinhua Qinggan Granules for Nonhospitalized COVID-19 Patients: a Double-Blind, Placebo-Controlled, Randomized Controlled Trial"

**Table S1.** Schedule of study assessments.

| **Study stage** | | **Screening** | **Medication** | **Follow up** |
| --- | --- | --- | --- | --- |
| Day(s) | | Day -5 to 0 | Day 1 | Day 10-15 |
| Signing informed consent forms | | √ |  |  |
| Determine inclusion / exclusion criteria | | √ | √ |  |
| Fill in the blanks with the general materials | | √ |  |  |
| Fill in medical history and treatment history | | √ |  |  |
| Concomitant diseases and medications | | √ | √ | √ |
| Physical Examination | | √ |  | √ |
| Signs & Symptoms | | √ |  | √ |
| Diagnostic indicators | | | | |
| Real-time PCR test | | √ |  | √ |
| Effectiveness  Indicators | Clinical Signs & Symptoms | √ | √ | √ |
|  | Real-time PCR test | √ |  | √ |
|  | Temperature | √ | √ | √ |
|  | Routine blood test | √ |  | √ |
|  | CRP test | √ |  | √ |
|  | Ferritin test | √ |  | √ |
|  | Lung X-ray examination | √ |  | √ |
|  | Quality of Life | √ |  | √ |
| Safety  Indicators | Basic vital signs | √ | √ | √ |
|  | Pulmonary signs | √ | √ | √ |
|  | Routine blood test | √ |  | √ |
|  | Urinalysis | √ |  | √ |
|  | Serum electrolytes | √ |  | √ |
|  | Liver function (ALT, AST, TBIL, AKP, γ-GT) | √ |  | √ |
|  | Renal function (BUN, Cr) | √ |  | √ |
|  | ECG | √ |  | √ |
|  | Adverse events |  |  | √ |
| Trial evaluation | | | | |
| Dispensation of quality-of-life assessment questionnaire | | √ |  | √ |
| Dispensation of investigational drug and diary card | |  | √ |  |
| Recycling quality of life assessment questionnaire | | √ |  | √ |
| Recycling of investigational drugs and diary card | |  |  | √ |
| Concomitant medication records | | √ | √ | √ |
| Analysis of reason for expulsion | |  |  | √ |
| Compliance evaluation | |  |  | √ |
| Safety evaluation | |  |  | √ |

**Table S2.** Seven-Category Ordinal Scale for Clinical Outcome

| **Clinical Status** | **Score** |
| --- | --- |
| Not hospitalized, able to resume normal activities; | 1 |
| Not hospitalized, but unable to resume normal activities; | 2 |
| Hospitalization, not requiring supplemental oxygen; | 3 |
| Hospitalization, requiring supplemental oxygen; | 4 |
| Hospitalization, requiring noninvasive mechanical ventilation; | 5 |
| Hospitalization, requiring invasive mechanical ventilation; | 6 |
| Death. | 7 |

**Table S3.** Biochemical indicators (SS)

| **Indicator** | **JHQG Group** | **Placebo Group** | **P-Value** |
| --- | --- | --- | --- |
| **Alanine aminotransferase** |  |  |  |
| Before treatment |  |  | 0.297 |
| N (N miss) | 150 (0) | 150 (0) |  |
| Mean (SD) | 31.33 (15.45) | 29.51 (14.71) |  |
| After treatment |  |  | 0.507 |
| N (N miss) | 129 (21) | 128 (22) |  |
| Mean (SD) | 30.31 (19.1) | 28.82 (16.73) |  |
| Before – after treatment |  |  | 0.981 |
| N (N miss) | 129 (21) | 128 (22) |  |
| Mean (SD) | -0.18 (13.97) | -0.22 (13.82) |  |
| Matching t-test | -0.145 /0.885 | -0.179 /0.858 |  |
| **Aspartate aminotransferase** |  |  |  |
| Before treatment |  |  | 0.108 |
| N (N miss) | 150 (0) | 150 (0) |  |
| Mean (SD) | 27.3667 (8.9775) | 25.8067 (7.7336) |  |
| After treatment |  |  | 0.088 |
| N (N miss) | 130 (20) | 128 (22) |  |
| Mean (SD) | 24.8615 (8.9614) | 23.2266 (6.1506) |  |
| Before treatment-After treatment |  |  | 0.750 |
| N (N miss) | 130 (20) | 128 (22) |  |
| Mean (SD) | 1.85 (8.97) | 2.16 (6.89) |  |
| Matching t-test | 2.347 /0.020 | 3.554 /0.001 |  |
| **Total Bilirubin** |  |  |  |
| Before treatment |  |  | 1.000 |
| N (N miss) | 150 (0) | 150 (0) |  |
| Mean (SD) | 0.5447 (0.2674) | 0.5447 (0.2898) |  |
| After treatment |  |  | 0.824 |
| N (N miss) | 129 (21) | 128 (22) |  |
| Mean (SD) | 0.58 (0.31) | 0.57 (0.32) |  |
| Before – after treatment |  |  | 0.527 |
| N (N miss) | 129 (21) | 128 (22) |  |
| Mean (SD) | -0.03 (0.25) | -0.05 (0.21) |  |
| Matching t-test | -1.609 /0.110 | -2.803 /0.006 |  |
| **Urea nitrogen** |  |  |  |
| Before treatment |  |  | 0.425 |
| N (N miss) | 149 (1) | 150 (0) |  |
| Mean (SD) | 10.0671 (3.2605) | 10.3533 (2.9290) |  |
| After treatment |  |  | 0.772 |
| N (N miss) | 130 (20) | 128 (22) |  |
| Mean (SD) | 9.5231 (3.1799) | 9.6328 (2.8968) |  |
| Before – after treatment |  |  | 0.412 |
| N (N miss) | 129 (21) | 128 (22) |  |
| Mean (SD) | 0.44 (2.72) | 0.73 (2.84) |  |
| Matching t-test | 1.848 /0.067 | 2.893 /0.004 |  |
| **Creatinine** |  |  |  |
| Before treatment |  |  | 0.711 |
| N (N miss) | 150 (0) | 150 (0) |  |
| Mean (SD) | 0.8169 (0.1495) | 0.8232 (0.1432) |  |
| After treatment |  |  | 0.317 |
| N (N miss) | 130 (20) | 128 (22) |  |
| Mean (SD) | 1.3432 (5.9817) | 0.8129 (0.1512) |  |
| Before-after treatment |  |  | 0.311 |
| N (N miss) | 130 (20) | 128 (22) |  |
| Mean (SD) | -0.53 (5.99) | 0.01 (0.11) |  |
| Matching t-test | -1.004 /0.317 | 0.988 /0.325 |  |
| **Alkaline phosphatase** |  |  |  |
| Before treatment |  |  | 0.239 |
| N (N miss) | 150 (0) | 150 (0) |  |
| Mean (SD) | 84.8667 (23.2995) | 89.8933 (46.6826) |  |
| After treatment |  |  | 0.670 |
| N (N miss) | 129 (21) | 128 (22) |  |
| Mean (SD) | 86.7829 (24.0419) | 88.1563 (27.4864) |  |
| Before – after treatment |  |  | 0.505 |
| N (N miss) | 129 (21) | 128 (22) |  |
| Mean (SD) | -1.98 (14.22) | -0.56 (19.35) |  |
| Matching t-test | -1.579 /0.117 | -0.329 /0.743 |  |
| **γ-transglutaminase** |  |  |  |
| Before treatment |  |  | 0.474 |
| N (N miss) | 150 (0) | 150 (0) |  |
| Mean (SD) | 43.5933 (36.6762) | 40.3333 (41.9963) |  |
| After treatment |  |  | 0.685 |
| N (N miss) | 129 (21) | 128 (22) |  |
| Mean (SD) | 39.2946 (42.6652) | 37.2969 (35.9961) |  |
| Before-after treatment |  |  | 0.451 |
| N (N miss) | 129 (21) | 128 (22) |  |
| Mean (SD) | 3.41 (18.03) | 0.93 (32.64) |  |
| Matching t-test | 2.148 /0.034 | 0.322 /0.748 |  |
| **Sodium** |  |  |  |
| Before treatment |  |  | 0.512 |
| N (N miss) | 150 (0) | 150 (0) |  |
| Mean (SD) | 139.0733 (2.2379) | 138.8933 (2.5014) |  |
| After treatment |  |  | 0.480 |
| N (N miss) | 130 (20) | 127 (23) |  |
| Mean (SD) | 138.9154 (2.1747) | 138.7165 (2.3296) |  |
| Before – after treatment |  |  | 0.954 |
| N (N miss) | 130 (20) | 127 (23) |  |
| Mean (SD) | 0.22 (2.73) | 0.24 (3.02) |  |
| Paired t-test | 0.901 /0.369 | 0.882 /0.380 |  |
| **Potassium** |  |  |  |
| Before treatment |  |  | 0.223 |
| N (N miss) | 150 (0) | 150 (0) |  |
| Mean (SD) | 4.2260 (0.4420) | 4.2840 (0.3781) |  |
| After treatment |  |  | 0.589 |
| N (N miss) | 130 (20) | 127 (23) |  |
| Mean (SD) | 4.2769 (0.3901) | 4.2528 (0.3244) |  |
| Before – after treatment |  |  | 0.090 |
| N (N miss) | 130 (20) | 127 (23) |  |
| Mean (SD) | -0.07 (0.52) | 0.03 (0.44) |  |
| Matching t-test | -1.616 /0.109 | 0.743 /0.459 |  |
| **Bicarbonate** |  |  |  |
| Before treatment |  |  | 0.169 |
| N (N miss) | 150 (0) | 150 (0) |  |
| Mean (SD) | 28.2520 (17.3978) | 26.2600 (3.2156) |  |
| After treatment |  |  | 0.904 |
| N (N miss) | 130 (20) | 127 (23) |  |
| Mean (SD) | 26.2491 (3.3287) | 26.3000 (3.4139) |  |
| Before -after treatment |  |  | 0.235 |
| N (N miss) | 130 (20) | 127 (23) |  |
| Mean (SD) | 2.21 (18.36) | 0.23 (3.71) |  |
| Paired t test | 1.370 /0.173 | 0.698 /0.487 |  |
| **Chloride** |  |  |  |
| Before treatment |  |  | 0.445 |
| N (N miss) | 150 (0) | 150 (0) |  |
| Mean (SD) | 103.7600 (7.1660) | 104.2467 (3.0848) |  |
| After treatment |  |  | 0.430 |
| N (N miss) | 130 (20) | 127 (23) |  |
| Mean (SD) | 104.4462 (2.6678) | 104.1969 (2.3706) |  |
| Before-after treatment |  |  | 0.306 |
| N (N miss) | 130 (20) | 127 (23) |  |
| Mean (SD) | -0.58 (7.87) | 0.18 (2.99) |  |
| Matching t-test | -0.846 /0.399 | 0.684 /0.495 |  |
| **Calcium** |  |  |  |
| Before treatment |  |  | 0.778 |
| N (N miss) | 150 (0) | 150 (0) |  |
| Mean (SD) | 9.1949 (0.4639) | 9.1799 (0.4573) |  |
| After treatment |  |  | 0.313 |
| N (N miss) | 129 (21) | 128 (22) |  |
| Mean (SD) | 10.0223 (7.6399) | 9.3386 (0.4437) |  |
| Before-after treatment |  |  | 0.333 |
| N (N miss) | 129 (21) | 128 (22) |  |
| Mean (SD) | -0.82 (7.61) | -0.17 (0.41) |  |
| Paired t-test | -1.226 /0.223 | -4.612 /<0.0001 |  |
| **Creatinine Kinase** |  |  |  |
| Before treatment |  |  | 0.640 |
| N (N miss) | 150 (0) | 150 (0) |  |
| Mean (SD) | 120.6467 (146.8939) | 113.7480 (104.9492) |  |
| After treatment |  |  | 0.307 |
| N (N miss) | 130 (20) | 128 (22) |  |
| Mean (SD) | 109.8615 (101.4561) | 99.4219 (55.6087) |  |
| Before-after treatment |  |  | 0.242 |
| N (N miss) | 130 (20) | 128 (22) |  |
| Mean (SD) | 2.60 (98.82) | 16.98 (98.22) |  |
| Matching t-test | 0.300 /0.765 | 1.956 /0.053 |  |

**Table S4.** Routine blood examination indicators (SS)

| **Indicators** | **JHQG Group** | **Placebo Group** | **P-Value** |
| --- | --- | --- | --- |
| **Red Blood Cell** |  |  |  |
| Before treatment |  |  | 0.655 |
| N (N miss) | 150 (0) | 150 (0) |  |
| Mean (SD) | 4.9099 (0.5902) | 4.8813 (0.5164) |  |
| After treatment |  |  | 0.955 |
| N (N miss) | 130 (20) | 128 (22) |  |
| Mean (SD) | 4.7804 (0.6135) | 4.7764 (0.5167) |  |
| Before - After treatment |  |  | 0.717 |
| N (N miss) | 130 (20) | 128 (22) |  |
| Mean (SD) | 0.11 (0.33) | 0.10 (0.25) |  |
| Matching t-test | 3.916 /0.000 | 4.593 /<0.0001 |  |
| **Haemoglobin** |  |  |  |
| Before treatment |  |  | 0.662 |
| N (N miss) | 150 (0) | 150 (0) |  |
| Mean (SD) | 13.8047 (1.6726) | 13.7193 (1.7025) |  |
| After treatment |  |  | 0.432 |
| N (N miss) | 130 (20) | 128 (22) |  |
| Mean (SD) | 13.5655 (1.6049) | 13.4031 (1.7092) |  |
| Before – after treatment |  |  | 0.773 |
| N (N miss) | 130 (20) | 128 (22) |  |
| Mean (SD) | 0.25 (0.66) | 0.27 (0.56) |  |
| Matching t-test | 4.280 /<0.0001 | 5.438 /<0.0001 |  |
| **White blood cell** |  |  |  |
| Before treatment |  |  | 0.738 |
| N (N miss) | 150 (0) | 150 (0) |  |
| Mean (SD) | 7.0640 (2.1717) | 6.9800 (2.1730) |  |
| After treatment |  |  | 0.579 |
| N (N miss) | 130 (20) | 128 (22) |  |
| Mean (SD) | 7.8885 (1.9038) | 7.7555 (1.9383) |  |
| Before -after treatment |  |  | 0.993 |
| N (N miss) | 130 (20) | 128 (22) |  |
| Mean (SD) | -0.80 (2.28) | -0.81 (2.09) |  |
| Matching t-test | -4.013 /0.000 | -4.363 /<0.0001 |  |
| **Neutrophil Count** |  |  |  |
| Before treatment |  |  | 0.644 |
| N (N miss) | 150 (0) | 150 (0) |  |
| Mean (SD) | 4.0468 (1.7613) | 3.9516 (1.8085) |  |
| After treatment |  |  | 0.948 |
| N (N miss) | 130 (20) | 128 (22) |  |
| Mean (SD) | 4.5347 (1.4942) | 4.5470 (1.5556) |  |
| Before-after treatment |  |  | 0.439 |
| N (N miss) | 130 (20) | 128 (22) |  |
| Mean (SD) | -0.46 (1.85) | -0.64 (1.91) |  |
| Matching t-test | -2.838 /0.005 | -3.802 /0.000 |  |
| **Neutrophil %** |  |  |  |
| Before treatment |  |  | 0.909 |
| N (N miss) | 150 (0) | 150 (0) |  |
| Mean (SD) | 55.9318 (10.8690) | 55.7920 (10.3176) |  |
| After treatment |  |  | 0.431 |
| N (N miss) | 130 (20) | 128 (22) |  |
| Mean (SD) | 57.0746 (8.1484) | 57.8844 (8.3312) |  |
| Before – after treatment |  |  | 0.439 |
| N (N miss) | 130 (20) | 128 (22) |  |
| Mean (SD) | -0.46 (1.85) | -0.64 (1.91) |  |
| Matching t-test | -2.838 /0.005 | -3.802 /0.000 |  |
| **Lymphocyte count** |  |  |  |
| Before treatment |  |  | 0.016 |
| N (N miss) | 150 (0) | 150 (0) |  |
| Mean (SD) | 2.1749 (0.6974) | 2.1461 (0.7467) |  |
| After treatment |  |  | 0.033 |
| N (N miss) | 130 (20) | 128 (22) |  |
| Mean (SD) | 2.3523 (0.6202) | 2.2771 (0.6256) |  |
| Before-after treatment |  |  | 0.049 |
| N (N miss) | 130 (20) | 128 (22) |  |
| Mean (SD) | -0.19 (0.58) | -0.12 (0.61) |  |
| Matching t-test | -3.653 /0.000 | -2.272 /0.025 |  |
| **Lymphocyte %** |  |  |  |
| Before treatment |  |  | 0.897 |
| N (N miss) | 150 (0) | 150 (0) |  |
| Mean (SD) | 32.1720 (10.1850) | 32.0273 (9.1190) |  |
| After treatment |  |  | 0.515 |
| N (N miss) | 130 (20) | 128 (22) |  |
| Mean (SD) | 30.6408 (7.0700) | 30.0704 (6.9736) |  |
| Before-after treatment |  |  | 0.378 |
| N (N miss) | 130 (20) | 128 (22) |  |
| Mean (SD) | 1.35 (8.00) | 2.25 (8.40) |  |
| Matching t-test | 1.926 /0.056 | 3.036 /0.003 |  |
| **Eosinophil%** |  |  |  |
| Before treatment |  |  | 0.927 |
| N (N miss) | 150 (0) | 150 (0) |  |
| Mean (SD) | 2.6860 (2.9480) | 2.7133 (2.1703) |  |
| After treatment |  |  | 0.103 |
| N (N miss) | 130 (20) | 128 (22) |  |
| Mean (SD) | 3.7162 (3.6938) | 3.1047 (2.0618) |  |
| Before – after treatment |  |  | 0.041 |
| N (N miss) | 130 (20) | 128 (22) |  |
| Mean (SD) | -0.95 (2.47) | -0.43 (1.49) |  |
| Matching t-test | -4.399 /<0.0001 | -3.259 /0.001 |  |
| **Blood platelets** |  |  |  |
| Before treatment |  |  | 0.295 |
| N (N miss) | 150 (0) | 150 (0) |  |
| Mean (SD) | 276.2600 (95.2808) | 265.5400 (81.0455) |  |
| After treatment |  |  | 0.989 |
| N (N miss) | 130 (20) | 128 (22) |  |
| Mean (SD) | 301.9000 (91.9734) | 302.0547 (93.2889) |  |
| Before -after treatment |  |  | 0.253 |
| N (N miss) | 130 (20) | 128 (22) |  |
| Mean (SD) | -24.75 (99.12) | -37.61 (80.20) |  |
| Matching t-test | -2.848 /0.005 | -5.305 /<0.0001 |  |

**Quality of Life Questionnaire (QOL)**

| Name: |  | Gender: |  | Age: |  | Drug No: |
| --- | --- | --- | --- | --- | --- | --- |

This questionnaire is designed to assess the impact of your disease (COVID-19) on various aspects of your life. This assessment asks how you feel about your quality of life, health, or other areas of your life. We ask what you think about your life in **the last one/two week(s).** Read each question carefully and FILL the circle forthe answer that best applies to you. Please answer ALL questions, as honestly as you can. About **the last one/two week(s)**, questions might ask:

| **Physiology Part** | **Before Treatment** | **After Treatment** |
| --- | --- | --- |
| **Questions** | **Day Zero** | **Day 10-15^th^** |
| Q1. How satisfied are you with your health?   ① Very dissatisfied ② Dissatisfied ③ Neither satisfied nor dissatisfied ④ Satisfied ⑤ Very satisfied |  |  |
| Q2. In the past one/two week(s), have you felt tired or fatigued due to your condition?   ① Not at all ② A little ③ A moderate amount ④ Very much ⑤ An extreme amount |  |  |
| Q3. To what extent do you feel that health status prevents you from doing what you need to do?   ① Not at all ② A little ③ A moderate amount ④ Very much ⑤ An extreme amount |  |  |
| Q4. Do you have enough energy for everyday life?   ① Not at all ② A little ③ Moderately ④ Mostly ⑤ Completely |  |  |
| Q5. In the last one/two week(s), have you experienced chest pains as a result of your cough?   ① Most of the time ② A good bit of the time ③ Some of the time  ④ A little of the time ⑤ None of the time |  |  |
| Q6. In the last one/two week(s), how much your condition disturbed your sleep?   ① Most of the time ② A good bit of the time ③ Some of the time   ④ A little of the time ⑤ None of the time |  |  |
| **Psychology Part** | **Before Treatment** | **After Treatment** |
| **Questions** | **Day Zero** | **Day 10-15^th^** |
| Q7. How would you rate your quality of life?   ① Very poor ② Poor ③ Neither poor nor good ④ Good ⑤ Very good |  |  |
| Q8. How much do you enjoy life?   ① Not at all ② A little ③ A moderate amount ④Very much ⑤ An extreme amount |  |  |
| Q9. In the last one/two week(s), have you felt any improvement of your condition?   ① Most of the time ② A good bit of the time ③ Some of the time ④ A little of the time ⑤ None of the time |  |  |
| Q10. How well are you able to concentrate?   ① Not at all ② A little ③ A moderate amount ④Very much ⑤ Extremely |  |  |
| Q11. In the last one/two week(s), have you worried that your condition may indicate serious illness?   ① Most of the time ② A good bit of the time ③ Some of the time ④ A little of the time ⑤ None of the time |  |  |
| Q12. How satisfied are you with your ability to perform your daily living activities?   ① Very dissatisfied ② Dissatisfied ③ Neither satisfied nor dissatisfied ④ Satisfied ⑤ Very satisfied |  |  |
| Q13. How satisfied are you with your capacity for work?   ① Very dissatisfied ② Dissatisfied ③ Neither satisfied nor dissatisfied ④ Satisfied ⑤ Very satisfied |  |  |
| Q14. How satisfied are you with yourself?   ① Very dissatisfied ② Dissatisfied ③ Neither satisfied nor dissatisfied ④ Satisfied ⑤ Very satisfied |  |  |
| Q15. How often do you have negative feelings (anxiety, depression) about your current health status?   ① Never ② Seldom ③ Quite often ④ Very often ⑤ Always |  |  |
| **Society** | **Before Treatment** | **After Treatment** |
| **Questions** | **Day Zero** | **Day 10-15^th^** |
| Q16. In the past one/two week(s), have you felt that due to your condition your family or friends have started avoiding you?   ① Not at all ② A little ③ A moderate amount ④Very much ⑤ An extreme amount |  |  |
| Q17. How healthy is your physical environment?   ① Not at all ② A little ③ A moderate amount ④Very much ⑤ Extremely |  |  |
| Q18. In the past one/two week(s), how much medical information is available to you about your current health status/condition?   ① Not at all ② A little ③ Moderately ④ Mostly ⑤Completely |  |  |
| Q19. To what extent do you have the opportunity for leisure activities?   ① Not at all ② A little ③ Moderately ④ Mostly ⑤Completely |  |  |
| Q20. How satisfied are you with the support you get from your friends?   ① Very dissatisfied ② Dissatisfied ③ Neither satisfied nor dissatisfied ④ Satisfied ⑤ Very satisfied |  |  |

**Marking procedure for QOL (The following should be filled by Physicians)**

| ***Field (Questions)*** | ***Marks Before Treatment*** | ***Marks After Treatment*** |
| --- | --- | --- |
|  | **Day Zero** | **Day 10-15^th^** |
| ① Physiology: Q1– Q6 | = ( ) ÷ 6 = | = ( ) ÷ 6 = |
| ② Psychology: Q7–Q15 | = ( ) ÷ 9 = | = ( ) ÷ 9 = |
| ③ Society: Q16 – Q20 | = ( ) ÷ 5 = | = ( ) ÷ 5 = |
| **Total score** |  |  |

***Note:***

Total score = {Sum of scores in three field (score: 3-21) (up to two decimal points)}

Note: Score in each field = Total score of each question ÷ Number of questions [(Score1-5) (up to two decimal points)]
